# Insulinopathies of the brain? Genetic overlap between somatic insulin-related and neuropsychiatric disorders

**DOI:** 10.1101/2021.05.28.21258000

**Authors:** Giuseppe Fanelli, Barbara Franke, Ward De Witte, I. Hyun Ruisch, Jan Haavik, Veerle van Gils, Willemijn J. Jansen, Stephanie J. B. Vos, Lars Lind, Jan K. Buitelaar, Tobias Banaschewski, Søren Dalsgaard, Alessandro Serretti, Nina Roth Mota, Geert Poelmans, Janita Bralten

## Abstract

The prevalence of Alzheimer’s disease (AD), autism spectrum disorder (ASD), and obsessive-compulsive disorder (OCD) is higher among patients with somatic insulinopathies, like metabolic syndrome (MetS), obesity, and type 2 diabetes mellitus (T2DM). Dysregulation of insulin signalling has been implicated in these neuropsychiatric disorders, and shared genetic factors might partly underlie these observed comorbidities. We investigated genetic overlap between AD, ASD, and OCD with MetS, obesity, and T2DM by estimating pairwise genetic correlations using the summary statistics of the largest available genome-wide association studies for these diseases. Stratified covariance analyses were performed to investigate the contribution of insulin-related gene-sets. Having tested these hypotheses, novel brain “insulinopathies” were explored by estimating the genetic relationship of six additional neuropsychiatric disorders with nine insulin-related diseases/traits. Significant genetic correlations were found between OCD and MetS (r_g_=-0.315, p=3.9e-8), OCD and obesity (r_g_=-0.379, p=3.4e-5), and OCD and T2DM (r_g_=-0.172, p=3e-4). Stratified analyses showed negative genetic covariances between ASD and MetS/T2DM through gene-sets comprising insulin signalling and/or insulin processing genes, and between AD/OCD and MetS/T2DM through an insulin receptor recycling gene-set (p<6.17e-4). Significant genetic correlations with insulin-related phenotypes were also found for anorexia nervosa, attention-deficit/hyperactivity disorder, major depression, and schizophrenia (p<6.17e-4). Our findings suggest the existence of two clusters of neuropsychiatric disorders, in which the genetics of insulin-related diseases and traits may exert divergent pleiotropic effects. These results represent a starting point for a new research line on “insulinopathies” of the brain, which may support the development of more effective/tolerated treatment strategies for neuropsychiatric disorders.

## Introduction

Mental disorders are characterised by a reduced life expectancy of approximately 10 years (1). In addition to violent causes of death, more than 67% of the increase in premature mortality is due to natural causes (2). The increased prevalence of insulin-related somatic diseases (i.e., type 2 diabetes mellitus (T2DM), obesity, and metabolic syndrome (MetS)) observed in mental disorders, with a resulting increased cardiovascular risk, contributes significantly to the lower life expectancy (3).

A number of studies have investigated this higher comorbidity, focusing mainly on metabolic disturbances as possible consequences of unhealthy lifestyles, sedentary habits, or the chronic use of psychotropic medication (4). However, there is growing evidence for the presence of glycaemic and metabolic imbalances in drug-naïve acute psychiatric patients already at disease onset, suggesting that common pathogenic mechanisms may also be involved (5). Shared genetic factors may play a role, and genomic studies may help to unravel the biological underpinnings that underlie the phenotypically observed comorbidity of neuropsychiatric disorders with somatic insulin-related diseases and traits. The above-mentioned insulin-related and neuropsychiatric diagnostic groups consist of complex and heterogeneous diseases with a highly polygenic inheritance pattern; heritability estimates from twin and family studies range between 30% and 80% (6, 7). Large meta-analyses of genome-wide association studies (GWASs) have identified hundreds of disease-associated single nucleotide polymorphisms (SNPs), each contributing with a small effect to the overall risk for these diseases (8). Genetic sharing has already been highlighted between T2DM, obesity and MetS, as expected from their highly interrelated pathogenesis (9), and recent evidence has also revealed the presence of substantial pleiotropy among psychiatric disorders (10).

A key feature that T2DM, obesity and MetS have in common is an impaired response to insulin stimulation in peripheral tissues, better known as insulin resistance (11). Abnormalities in insulin signalling might also link with neuropsychiatric disorders. Indeed, beyond the anabolic function of insulin at the peripheral level, where it promotes the glucose uptake in tissues while stimulating glycogenesis and lipogenesis, this hormone can also bind to insulin receptors (INSRs) on the surface of both neurons and glial cells in the central nervous system (CNS) (11), where insulin signalling is regulated a.o. by the neurotransmitters serotonin and dopamine (12). In the CNS, insulin plays a key role in synaptic plasticity and neurotransmission, apoptosis inhibition, and neuroinflammation (13). Preclinical studies have suggested that an increase in the mammalian target of rapamycin (mTOR) activity, one of the major downstream effectors of the INSRs, may lead to deficits in synaptic pruning, and thereby contributes to the cognitive inflexibility and perseverative/repetitive behaviours observed in those animals with mTOR genetic alterations (14, 15). Cognitive abnormalities of a similar nature were shown in TALLYHO/JngJ mice, an animal model of T2DM (16).

Recently, dysregulation in insulin signalling has been suggested to contribute to neuropsychiatric disorders characterised by cognitive inflexibility more widely. Evidence is strongest for Alzheimer’s disease (AD) and autism spectrum disorder (ASD) (17-22). Our own recent work also suggested a link with obsessive-compulsive disorder (OCD) (18, 22). In the case of AD, it has been shown that insulin sensitivity is altered even before the onset of cognitive decline or β-amyloid (Aβ) accumulation in the CNS (20). The hyperactivation of the phosphatidylinositol-3-kinase (PI3K)/protein kinase B (AKT)/mTOR cascade, mediated by the phosphorylation of INSR via insulin binding to the neuronal surface, leads to the inhibition of autophagy processes and subsequent accumulation of damaged mitochondria and misfolded proteins seen in AD (19). The same PI3K/AKT/mTOR hyperactivation is also involved in ASD pathogenesis (17), and genes within the mTOR pathway were also shown to associate with brain volume variability and ASD (Arenella et al. 2021, submitted). Furthermore, offspring of mothers who have T2DM during pregnancy have a higher risk of developing ASD (21). For OCD the evidence for alterations in CNS insulin signalling comes from the integration of data from different types of genetic studies, and this potential association may drive alterations in excitatory synaptogenesis and dendritic spine formation (18). Also obsessive-compulsive symptoms in the general population have been associated with CNS insulin signalling (22), and shared genetic aetiologies of peripheral insulin-related phenotypes (i.e., T2DM, glucose levels 2 hours after an oral glucose challenge (2hGlu), and fasting insulin (FIns)) were found with both obsessive-compulsive symptoms and OCD (22). In light of the above evidence, we aimed to investigate the extent of the potential genetic sharing and contribution of insulin-related gene-sets in the observed comorbidity of neuropsychiatric disorders characterised by cognitive inflexibility (i.e., AD, ASD, and OCD) with somatic diseases related to insulin resistance, namely MetS, obesity, and T2DM. For this purpose, we performed Linkage Disequilibrium SCore regression (LDSC) and stratified GeNetic cOVariance Analyzer (GNOVA) analyses (23, 24). In addition, we explored potential additional brain “insulinopathies” by estimating the genetic overlap between other neuropsychiatric disorders and insulin-related somatic phenotypes.

## Materials and methods

### Input datasets

As input for the analyses, we used summary statistic data of the largest GWASs available at the time of conducting our analyses for the phenotypes of interest (see also Table 1 and the Supplementary information). We considered the most prevalent somatic diseases linked to insulin-resistance (i.e., MetS, obesity, and T2DM), and neuropsychiatric disorders linked to cognitive inflexibility, namely AD, ASD, and OCD. We also investigated insulin-related traits (i.e., 2hGlu, body mass index (BMI), fasting glucose (FGlu) and FIns, glycated haemoglobin (HbA1c), and homeostatic model assessment for insulin resistance (HOMA-IR)), and other six neuropsychiatric disorders, which are those best characterised genetically by the Psychiatric Genomic Consortium (10) (i.e., attention-deficit hyperactivity disorder (ADHD), anorexia nervosa (AN), bipolar disorder (BIP), major depressive disorder (MDD), schizophrenia (SCZ), and Tourette’s syndrome (TS)). Data were downloaded from online repositories (see <u>URLs</u>), when publicly available, or requested (i.e., MetS) from the GWAS authors.

**Table 1.** Characteristics of the samples used for the linkage-disequilibrium score regression (LDSC) and GeNetic cOVariance Analyzer (GNOVA) analyses. Abbreviations: 2hGlu, glucose levels 2 hours after an oral glucose challenge; BMI, body mass index; FGlu, fasting glucose; FIns, fasting insulin; HbA1c, glycated haemoglobin; HOMA-IR, homeostatic model assessment for insulin resistance; MetS, metabolic syndrome; T2DM, type 2 diabetes mellitus; ADHD, attention-deficit/hyperactivity disorder; AD, Alzheimer’s disease; AN, anorexia nervosa; ASD, autism spectrum disorder; BIP, bipolar disorder; MDD, major depressive disorder; OCD, obsessive-compulsive disorder; SCZ, schizophrenia; TS, Tourette’s syndrome; N, total sample size; Neff, effective sample size [Neff = 4 / (1/Cases + 1/Controls)].

| Trait/Disorder | Author | Year | PMID | Consortium | Ancestry | N | Cases | Controls | Neff |
| --- | --- | --- | --- | --- | --- | --- | --- | --- | --- |
| **2hGlu** | Saxena et al. | 2010 | 20081857 | MAGIC | European | 15,234 |  |  |  |
| **BMI** | Pulit et al. | 2018 | 30239722 | GIANT | European | 697,734 |  |  |  |
| **FGlu** | Lagou et al. | 2021 | 33402679 | MAGIC | European | 140,595 |  |  |  |
| **FIns** | Lagou et al. | 2021 | 33402679 | MAGIC | European | 98,210 |  |  |  |
| **HbA1c** | Wheeler et al. | 2017 | 28898252 | MAGIC | European | 123,665 |  |  |  |
| **HOMA-IR** | Dupuis et al. | 2010 | 20081858 | MAGIC | European | 37,037 |  |  |  |
| **MetS** | Lind | 2019 | 31589552 |  | European | 291,107 | 59,677 | 231,430 | 189,772.64 |
| **Obesity** | Watanabe et al. | 2019 | 31427789 |  | European | 244,890 | 9,805 | 235,085 | 37,649.69 |
| **T2DM** | Mahajan et al. | 2018 | 30297969 | DIAGRAM | European | 898,130 | 74,124 | 824,006 | 272,025.75 |
| **ADHD** | Demontis et al. | 2019 | 30478444 | PGC | European | 53,293 | 19,099 | 34,194 | 49,017.41 |
| **AD** | Jansen et al. | 2019 | 30617256 |  | European | 455,258 | 71,880 | 383,378 | 242,123.90 |
| **AN** | Watson et al. | 2019 | 31308545 | PGC | European | 72,517 | 16,992 | 55,525 | 52,041.91 |
| **ASD** | Grove et al. | 2019 | 30804558 | PGC | European | 46,350 | 18,381 | 27,969 | 44,366.62 |
| **BIP** | Stahl et al. | 2018 | 31043756 | PGC | European | 51,710 | 20,352 | 31,358 | 49,367.47 |
| **OCD** | OC GAS/IOCDF-GC | 2018 | 28761083 | OC GAS/IOCDF-GC | European | 9,725 | 2,688 | 7,037 | 7,780.14 |
| **MDD** | Wray et al./<br>Howard et al. | 2019 | 29700475/<br>29662059 | PGC | European | 500,199 | 170,756 | 329,443 | 449,855.91 |
| **SCZ** | Pardinas et al. | 2018 | 29483656 | PGC+CLOZUK | European | 105,318 | 40,675 | 64,643 | 99,863.42 |
| **TS** | Yu et al. | 2019 | 30818990 | PGC | European | 14,307 | 4,819 | 9,488 | 12,783.30 |

### Genome-wide bivariate genetic correlation estimations

Bivariate LDSC (https://github.com/bulik/ldsc) analyses were performed to estimate the genetic correlation (r_g_) ascribed genome-wide to common variants between AD, ASD, OCD, and MetS, obesity, and T2DM, following the software guidelines (https://github.com/bulik/ldsc/wiki/Heritability-and-Genetic-Correlation). Also through LDSC, exploratory analyses were carried out to estimate the extent of the genetic sharing between other neuropsychiatric disorders (ADHD, AN, BIP, MDD, SCZ, TS, along with AD, ASD, and OCD) and insulin-related somatic diseases/traits (i.e., 2hGlu, BMI, FGlu and FIns, HbA1c, HOMA-IR, along with MetS, obesity, and T2DM). Further details on the quality control (QC) steps and the LDSC method are provided in the Supplementary information. LDSC is computationally robust to sample overlaps between studies (23). Bonferroni correction was applied, accounting for the number of analyses performed (α=0.05/(9* 9)=6.17e-4).

### Genetic covariance analyses stratified by functional annotations

GNOVA (https://github.com/xtonyjiang/GNOVA) was used to investigate whether AD, ASD, and OCD were genetically correlated to MetS, obesity, or T2DM specifically through nine gene-sets involved in peripheral and/or CNS insulin signalling (gene-set sizes ranged from 27 to 137 genes; see Table S4-S5 for a complete list of genes included in each gene-set). Further details on the GNOVA method and the selection of the insulin signalling-related gene-sets are provided in the Supplementary information. GNOVA-computed covariance estimates are robust to sample overlaps (24). Bonferroni correction was applied to GNOVA results considering the nine tested gene-sets and the nine combinations of insulin-related somatic diseases and neuropsychiatric disorders for which GNOVA analyses were performed (α=0.05/(9* 9)=6.17e-4).

## Results

### Description of the input datasets

A description of the samples (with sample sizes, number of cases and controls, and the derived effective sample size) included in the analyses is provided in Table 1. Further information on the GWAS samples can be found in the Supplementary information.

### Pairwise genome-wide genetic correlations between neuropsychiatric disorders characterised by cognitive inflexibility and insulin-related somatic diseases and traits

A genetic correlation plot depicting the LDSC analyses results is shown in Figure 1; details on the genetic correlation estimates (r_g_) for each pair and statistical significance are provided in Table 2. After correcting for multiple testing, negative genetic correlations were highlighted between OCD and MetS (r_g_=-0.315, p=3.9e-8), OCD and obesity (r_g_=-0.379, p=3.6e-5), and OCD and T2DM (r_g_=-0.172, p=3e-4). Nominally significant genetic correlations were also found between AD and MetS (r_g_=0.166, p=0.018), AD and T2DM (r_g_=0.175, p=0.013), and ASD and MetS (r_g_=0.115, p=0.002).

**Table 2.** Genetic correlation table reporting the detailed results derived from the bivariate Linkage Disequilibrium Score regression (LDSC) analyses. Reported values are genetic correlation estimates – r_g_ – (p-values). Abbreviations: AD, Alzheimer’s disease; ASD, autism spectrum disorders; OCD, obsessive-compulsive disorder; ADHD, attention-deficit/hyperactivity disorder; AN, anorexia nervosa; BIP, bipolar disorder; MDD, major depressive disorder; SCZ, schizophrenia; TS, Tourette’s syndrome; MetS, metabolic syndrome; T2DM, type 2 diabetes mellitus; 2hGlu, glucose levels 2 hours after an oral glucose challenge; BMI, body mass index; FGlu, fasting glucose; FIns, fasting insulin; HbA1c, glycated haemoglobin; HOMA-IR, homeostatic model assessment for insulin resistance. * * Statistically significant bivariate genetic correlation (p<6.17e-4) * Nominally significant bivariate genetic correlation (p<0.05)

| Trait/Disorder | AD | ASD | OCD | ADHD | AN | BIP | MDD | SCZ | TS |
| --- | --- | --- | --- | --- | --- | --- | --- | --- | --- |
| **MetS** | 0.166 (0.018) * | 0.115 (0.002) ** | -0.315 (3.88e-8) ** | 0.386 (7.16e-30) ** | -0.279 (3.43e-15) ** | -0.027 (0.349) | 0.177 (1.66e-16) ** | -0.090 (1.41e-5) ** | -0.026 (0.496) |
| **Obesity** | 0.134 (0.188) | 0.115 (0.072) | -0.379 (3.35e-5) ** | 0.538 (9.91e-24) ** | -0.250 (7.60e-6) ** | -0.017 (0.716) | 0.235 (5.31e-10) ** | -0.087 (0.009) * | 0.042 (0.552) |
| **T2DM** | 0.175 (0.013) * | 0.035 (0.403) | -0.172 (3e-4) ** | 0.328 (3.24e-28) ** | -0.209 (6.04e-12) ** | -0.049 (0.057) | 0.141 (4.65e-11) ** | -0.044 (0.016) * | 0.013 (0.713) |
| **2hGlu** | 0.137 (0.409) | 0.009 (0.936) | 0.090 (0.559) | -0.004 (0.964) | -0.122 (0.221) | 0.086 (0.344) | 0.006 (0.927) | -0.020 (0.743) | -0.112 (0.495) |
| **BMI** | 0.159 (0.007) * | 0.043 (0.164) | -0.284 (2.57e-11) ** | 0.348 (6.59e-49) ** | -0.308 (6.38e-38) ** | -0.063 (0.007) * | 0.112 (1.55e-10) | -0.017 (7.95e-11) ** | -0.008 (0.801) |
| **FGLu** | 0.030 (0.630) | -0.043 (0.334) | -0.072 (0.339) | 0.123 (6e-4) ** | -0.126 (0.005) * | -0.012 (0.740) | 0.070 (0.012) * | -0.027 (0.296) | 0.089 (0.074) |
| **Flns** | 0.067 (0.483) | -0.017 (0.797) * | -0.108 (0.198) | 0.154 (0.005) * | -0.303 (4.17e-7) ** | -0.060 (0.203) | 0.088 (0.045) * | -0.029 (0.464) | 0.089 (0.190) |
| **HbA1C** | -0.081 (0.241) | -0.030 (0.619) | 0.079 (0.367) | 0.124 (0.006) * | -0.155 (0.003) * | -0.012 (0.769) | 0.012 (0.722) | -0.009 (0.751) | 0.075 (0.225) |
| **HOMA-IR** | -0.1132 (0.3215) | 0.026 (0.817) | -0.139 (0.255) | 0.1313 (0.138) | -0.3029 (1e-4) ** | -0.033 (0.616) | 0.1044 (0.076) | -0.0278 (0.577) | 0.086 (0.390) |

**Figure 1.**
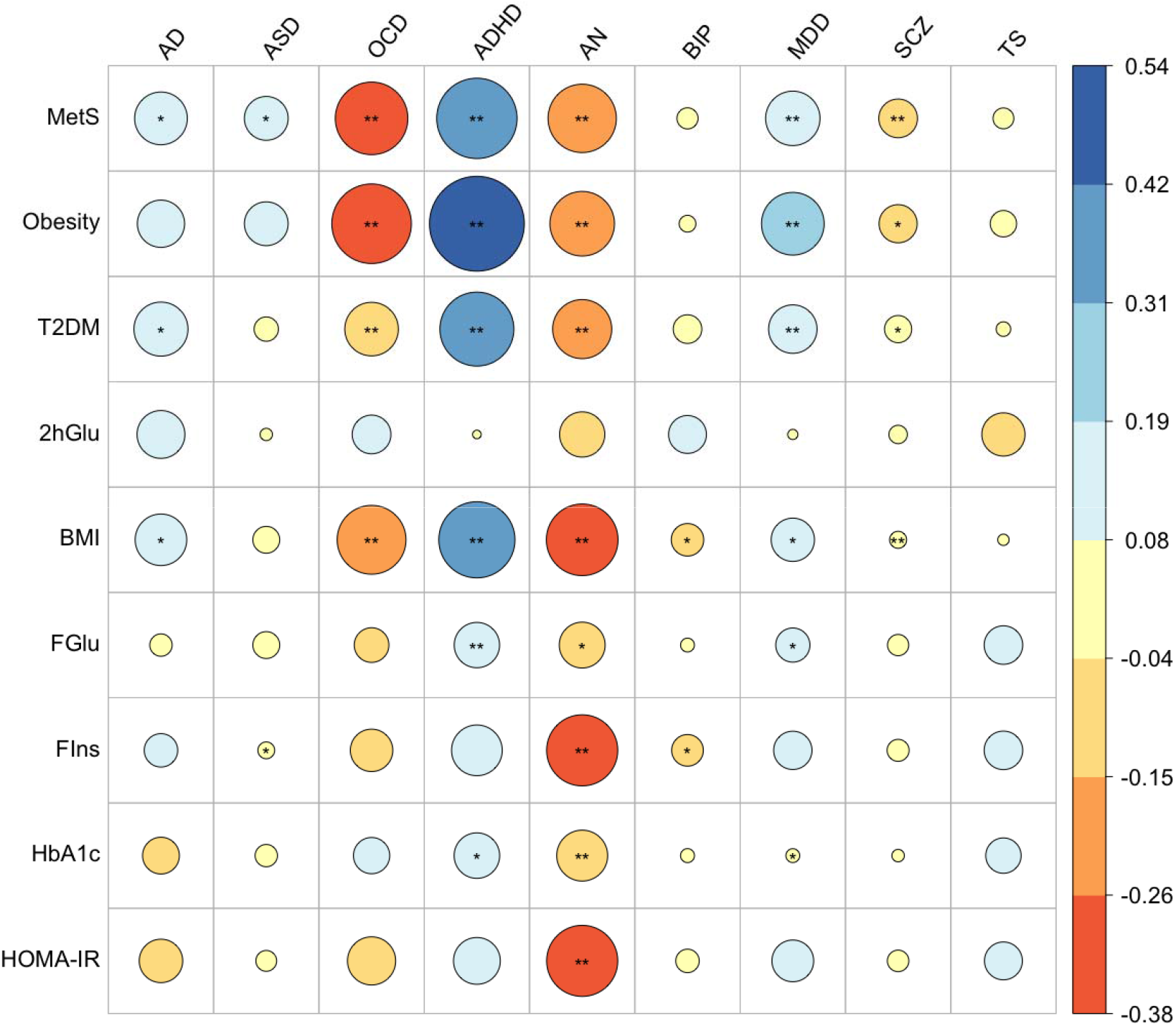
Genetic correlation plot summarising the results from the bivariate Linkage Disequilibrium Score regression (LDSC) analyses. The size of the circle is proportional to the genetic correlation estimates, going from colder to warmer colours as the direction of the effect changes from positive to negative. Bonferroni multiple testing correction was applied, correcting for the number of analyses performed (α=0.05/(9* 9)=6.17e-4). Abbreviations: AD, Alzheimer’s disease; ASD, autism spectrum disorder; OCD, obsessive-compulsive disorder; ADHD, attention-deficit/hyperactivity disorder; AN, anorexia nervosa; BIP, bipolar disorder; MDD, major depressive disorder; SCZ, schizophrenia; TS, Tourette’s syndrome; MetS, metabolic syndrome; T2DM, type 2 diabetes mellitus; 2hGlu, glucose levels 2 hours after an oral glucose challenge; BMI, body mass index; FGlu, fasting glucose; FIns, fasting insulin; HbA1c, glycated haemoglobin; HOMA-IR, homeostatic model assessment for insulin resistance. *Nominally significant bivariate genetic correlation (p <0.05) * * Statistically significant bivariate genetic correlation (p<6.17e-4)

When insulin-related somatic traits (i.e., 2hGlu, BMI, FGlu, FIns, HbA1c, HOMA-IR) were considered, OCD was also found to be significantly negatively genetically correlated with BMI (r_g_=-0.284, p=2.57e-11), but neither AD nor ASD showed significant correlations with the traits.

### Genetic covariance of AD, ASD, and OCD with MetS, obesity and T2DM stratified by insulin-related gene-sets

After Bonferroni correction, stratified GNOVA analyses highlighted significant negative genetic covariance between AD and MetS through the Reactome INSR recycling gene-set (ρ_g_=-2e-4, r_g_=-0.82, p=1e-5), between ASD and T2DM through the Reactome insulin processing gene-set (ρ_g_=-5e-4, r_g_=-0.45, p=2.8e-4), as well as between ASD and MetS through the Biocarta, KEGG, and PID insulin signalling pathways (ρ_g_=-4.1e-4, r_g_=-0.73, p=2e-5; ρ_g_=-0.002, r_g_=-0.73, p=3e-5; ρ_g_=-8e-4, p=1e-5, respectively). OCD showed negative genetic covariance with MetS and T2DM through the Reactome INSR recycling gene-set (ρ_g_=-0.001, r_g_=-0.81, p=1e-5, and ρ_g_=-0.001, r_g_=-0.51, p=1.6e-4, respectively). No genetic covariance between AD, ASD, OCD and obesity was found at the gene-sets level (Table 3; detailed results are shown in Tables S1-S3).

**Table 3.** Summary results of the genetic covariance analyses between AD, ASD, OCD and somatic diseases linked with insulin-resistance stratified by insulin signalling gene-sets. Abbreviations: AD, Alzheimer’s disease; ASD, autism spectrum disorder; OCD, obsessive-compulsive disorder; MetS, metabolic syndrome; T2DM, type 2 diabetes mellitus. * * Statistically significant stratified genetic covariance estimates (p<6.17e-4) ^a^ ρ_g_ corrected: genetic covariance estimates with sample overlap correction ^b^ p corrected: p-value from the statistical test for genetic covariance with sample overlap correction ^c^ r_g_ corrected: genetic correlation estimates with sample overlap correction § the noise in the estimation of single-nucleotide polymorphism (SNP)-based heritability may cause the genetic correlation estimates to be out of bounds (>1) or even be set as missing (NA) when the SNP-based heritability estimates are small or below zero, respectively. In such cases, genetic covariance estimates are less biased than genetic correlation measures. ρ_g_: genetic covariance estimate SE ρ_g_: standard error of the estimate of ρ_g_ p: p-value from the statistical test for genetic covariance r_g_: genetic correlation estimate h^2^_SNP_ 1: SNP-based heritability estimate for the first phenotype h^2^_SNP_ 2: SNP-based heritability estimate for the second phenotype

| Gene-set name | n genes/<br>gene-set | Base<br>phenotypes | $\rho_g$ | SE $\rho_g$ | $p$ | $r_g$ | $h^2_{\text{SNP } 1}$ | $h^2_{\text{SNP } 2}$ | annotated<br>SNPs | total SNPs |
| --- | --- | --- | --- | --- | --- | --- | --- | --- | --- | --- |
| REACTOME INSULIN RECEPTOR RECYCLING | 26 | AD x MetS | -0.00022 <sup>a</sup> | 0.00005 | $1 \times 10^{-5}$ <sup>b</sup> | -0.82 | 0.00010 | 0.00074 | 2,132 | 1,019,413 |
| BIOCARTA INSULIN PATHWAY | 21 | ASD x MetS | -0.00041 | 0.00010 | $2 \times 10^{-5}$ | -0.73 | 0.00046 | 0.00068 | 1,520 | 968,964 |
| KEGG INSULIN SIGNALING PATHWAY | 137 | ASD x MetS | -0.00170 | 0.00041 | $3 \times 10^{-5}$ | -0.73 | 0.00261 | 0.00207 | 11,334 | 968,964 |
| PID INSULIN PATHWAY | 44 | ASD x MetS | -0.00080 | 0.00018 | $1 \times 10^{-5}$ | -2.25 § | 0.00012 | 0.00105 | 4,319 | 968,964 |
| REACTOME INSULIN PROCESSING | 27 | ASD x T2DM | -0.00050 | 0.00014 | $3 \times 10^{-4}$ | -0.45 | 0.00090 | 0.00137 | 3,094 | 968,306 |
| REACTOME INSULIN RECEPTOR RECYCLING | 26 | OCD x MetS | -0.00124 | 0.00028 | $1 \times 10^{-5}$ | -0.81 | 0.00316 | 0.00074 | 2,132 | 1,019,413 |
| REACTOME INSULIN RECEPTOR RECYCLING | 26 | OCD x T2DM | -0.00100 | 0.00026 | $2 \times 10^{-4}$ | -0.51 | 0.00304 | 0.00128 | 2,130 | 1,019,648 |

### In search of new brain “insulinopathies”: LDSC-based genetic correlation analyses for additional neuropsychiatric disorders and insulin-related diseases and traits

Analyses were also extended to other neuropsychiatric disorders (i.e., ADHD, AN, BIP, MDD, SCZ, and TS). Significant genetic correlations between the aforementioned disorders and insulin-related diseases/traits were found for ADHD, AN, MDD, and SCZ (see Figure 1 and Table 2).

## Discussion

In this study, we investigated the genetic links between neuropsychiatric disorders featuring cognitive inflexibility (i.e., AD, ASD, OCD) and somatic insulinopathies, namely MetS, obesity and T2DM, hypothesising an important role for genes related to insulin signalling. Our genome-wide analyses indicate significant genetic correlations between OCD and obesity, T2DM, and MetS. Gene-set stratified genetic covariance analyses of specific insulin-related pathways helped identify a genetic link of AD, ASD, and OCD with MetS, and a link of ASD and OCD with T2DM. Our exploration of potentially novel brain “insulinopathies” yielded evidence for genetic overlap of ADHD, AN, MDD, and SCZ with diseases/traits linked with insulin resistance.

A role of metabolic dysregulation in ASD has been previously suggested by the increased risk for ASD and neurodevelopmental delays in the offspring of mothers who have metabolic conditions during pregnancy (25). Nevertheless, our study did not find ASD to be significantly genetically correlated at the genome-wide level with either MetS, obesity or T2DM, in line with previous reports using smaller sample sizes (26). However, the stratification to insulin-specific gene-sets revealed significant localised negative genetic covariance of ASD with MetS and T2DM through genes within insulin processing and signalling pathways. Although further studies will be needed to disentangle the biological meaning of these findings, we could speculate that the observed pathway-level negative genetic correlation between ASD and MetS/T2DM might reflect higher complexity of reciprocal regulation between monoamine and insulin signalling at the CNS and peripheral level (12). Interestingly, while the insulin processing pathway is generally involved in the generation and subsequent exocytosis of insulin-containing granules, individual genes within this set (i.e., syntaxin-1A (STX1A), vesicle-associated membrane protein 2/synaptobrevin-2 (VAMP2), proprotein convertase subtilisin/kexin type 2 (PCSK2)) have also been linked to presynaptic neurotransmission and previously been associated with autism, intellectual and neurodevelopmental disabilities (27, 28).

Few clinical and epidemiological studies are available providing information on the phenotypic overlap between OCD and somatic insulin-related diseases. The existing reports indicate a higher prevalence of obesity, MetS, and T2DM in patients with OCD than the general population (29, 30). Furthermore, a mouse model for T2DM showed compulsive traits, as discussed above (20). We thus had hypothesised a genetic correlation between OCD and somatic disorders characterised by insulin resistance to exist, which we indeed found in this study. The negative direction of the correlation we found was unexpected, as it might suggest a protective role of the genetics underlying OCD on the chance of having T2DM, MetS and/or obesity. However, for behavioural traits, environmental sources of variation may operate orthogonally to genetic factors, masking the effect of the genetics at the phenotypic level (31). Therefore, one hypothesis explaining our result can be that environmental effects act in the opposite direction to genetics, causing an increased risk in the presence of protective genetics and resulting in variability in the phenotypic manifestations over time. Indeed, metabolic complications have been particularly associated with a longer duration of antipsychotics treatment in patients with OCD (29). It is also reasonable to assume that patients with more severe symptoms, having higher genetic load for OCD, are more likely to develop metabolic side effects of such treatments because they require higher doses and longer therapies, even though they might be genetically more protected against insulin-related/metabolic disturbances. The analyses considering insulin-related glycaemic/anthropometric traits also showed a negative correlation between OCD and BMI. This finding is consistent with previous evidence in smaller samples of a negative genetic relationship between OCD and body fat measures (32); it also further supports the negative correlation trend that we observed between OCD and somatic insulinopathies. Zooming in through analyses of gene-sets related to insulin signalling, we found genes involved in the INSR recycling process involved in the genetic correlation of OCD with both MetS and T2DM. This molecular pathway mediates the recycling of the INSR and reintegration into the plasma membrane. After activation, the INSR-insulin complex is internalised into the cell within an endosome, and insulin is degraded, while INSR is dephosphorylated and reintegrated into the plasma membrane (33). To our knowledge, this is the first study reporting involvement of the INSR recycling pathway in neuropsychiatric phenotypes. In this respect, it should be noted that endosomal recycling processes are relevant to the functioning of the brain. They are important for synaptic functioning and plasticity (and related glutamatergic neurotransmission) as well as for the maintenance of levels of membrane proteins, more generally (34).

We did not observe significant genome-wide genetic correlations between AD and somatic insulin-related diseases, only nominally significant positive genetic correlations were seen with MetS and T2DM before multiple testing correction. Interactions between genes, genes and environment, and/or epigenetic effects may explain the comorbidity observed at the phenotypic level better than the additive genetic effects of SNPs explored by our strategy. It should also be considered that the risk for both AD and T2DM increases enormously with ageing. Processes linked to oxidative damage and ageing could trigger the onset of both diseases in a way that is partly independent from genetic effects (19). Another hypothesis is that the clinical heterogeneity of AD may have camouflaged the presence of genetic factors shared with somatic insulinopathies and that more deeply phenotyped samples may help better investigate the presence of pleiotropic effects (35). Alternatively or in addition, previous evidence has also shown a strong regional genetic correlation between AD and T2DM for the genetic variants mapped to the apolipoprotein-E (*APOE*) locus (36). Furthermore, oral antidiabetics, such as thiazolidinediones, and intranasal insulin have shown differential efficacy in AD depending on the *APOE*-ε4 genotype (37), which is the strongest common genetic risk factor for late-onset AD (38). This previous evidence may suggest a role for insulin signalling specifically in individuals carrying *APOE* polymorphisms, suggesting that new insights may be derived from stratification of the AD population according to *APOE* genotype. The absence of genetic correlations at the genome-wide level does not preclude the existence of genetic sharing, as both positive and negative local genetic correlations may occur and potentially cancel each other out when summed at the genome-wide level (39). In this regard, we demonstrated significant genetic covariance between AD and MetS at the INSR recycling gene-set level. Under physiological conditions, INSR is maintained in equilibrium between an internalising and an exposed state at the plasma membrane (40). Either excessive or insufficient surface INSR can lead to the development of insulin resistance (40). Our finding is in line with the evidence of an altered cellular distribution of INSRs in AD, resulting in a loss of INSRs at the neuronal membrane, suggesting that alterations in INSR recycling/trafficking are present (41).

Interestingly, the local genetic covariance we have highlighted between neuropsychiatric disorders characterised by reduced cognitive flexibility and somatic diseases linked to insulin resistance was in all cases in the negative direction at the level of gene-sets related to insulin signalling. This means that genetic variability at the level of these gene-sets may result in an opposite pleiotropic effect on these two groups of diseases. However, the biological interpretation of these findings does not seem obvious at present and additional investigations at the gene and functional level will be necessary to clarify their biological significance.

To extend the spectrum of potential brain “insulinopathies”, LDSC analyses were performed considering six other neuropsychiatric disorders and diseases/traits related to insulin resistance. Our analyses identified several additional genetic correlations of the somatic insulin-related diseases with psychiatric disorders; negative genetic correlations were seen between MetS and both AN and schizophrenia, and positive genetic correlations were observed for MetS with both ADHD and MDD. The diagnosis of MetS is made when at least three out of the following co-occur: high systolic blood pressure, low levels of high-density lipoprotein (HDL), hyperglycaemia, high levels of triglycerides, and/or increased waist circumference (9). Our findings are consistent with previous evidence of pairwise genetic sharing between lipidemic traits (HDL and triglycerides), waist circumference and AN, ADHD, and/or MDD (8, 42-44). In line with the negative genetic correlations that we observed between MetS and both AN and schizophrenia, MR studies have previously shown a causal relationship between decreased fat mass and both AN and SCZ (32). This finding may suggest a prevalent contribution of environmental factors, such as the use of antipsychotics, unhealthy diet and lifestyle, reduced access to medical care on the epidemiological evidence of an increased risk of MetS, hypertension, and dyslipidaemia in patients with SCZ (45). We also replicated and updated previous evidence of genetic sharing of ADHD, AN, and MDD with T2DM, as well as of ADHD, AN, MDD, and SCZ with both obesity and BMI (8, 23, 32, 42, 43, 46). With regard to SCZ and BMI, the negative direction of the genetic correlation corresponds to the previously reported evidence of a negative association of polygenic risk scores for SCZ with BMI (46). Exploring further the genetic links between these neuropsychiatric disorders and glycaemic traits linked to insulin resistance, we revealed a novel positive correlation between ADHD and FGlu, as well as negative bivariate correlations between AN and both FIns and HOMA-IR that replicate and update previous findings (32, 42). A Mendelian randomisation study had also previously shown that higher levels of FIns have a causal effect in reducing the risk of AN (47).

This study comes with some strengths and limitations. The major strength is the investigation of the possible specific involvement of insulin-related gene-sets at the genomic level for the first time in the phenotypically observed comorbidity between neuropsychiatric diseases related to cognitive inflexibility (i.e., AD, ASD, OCD) and somatic diseases related to insulin resistance. GNOVA provided us with more powerful statistical inference and more accurate genetic covariance estimates than LDSC and helped dissect the shared genetic architecture of the considered complex diseases while giving us greater insights into the underlying biology. We exploited the largest public GWAS summary statistics (up to 898,130 individuals for T2DM) and used a strict Bonferroni correction to avoid type-1 errors. Our study may be limited by not having considered in our analyses the potential effect of environmental factors and epigenetic mechanisms, which are likely to mediate the relationship between neuropsychiatric and somatic insulinopathies. In addition, the composition of insulin-related gene-sets, used as functional annotations in our stratified analyses, may be influenced by the current, still incomplete knowledge of the biology and functioning of the pathways to which they refer.

In conclusion, our study revealed the presence of genetic overlap between OCD and insulin-related somatic diseases, with a likely protective effect of the genetics underlying OCD on the chance of having MetS, obesity, and/or T2DM. However, environmental effects, such as psychotropic drug use, or a relatively unhealthy lifestyle, may act in the opposite direction to genetics, causing increased metabolic risk despite protective genetics. We further demonstrated that insulin-related gene-sets may be pleiotropic for neuropsychiatric disorders related to cognitive inflexibility and MetS or T2DM, in all cases suggesting that the cumulative effect of genetic variability within insulin-related gene-sets on the neuropsychiatric disorders investigated is in the opposite direction to the effect on MetS and T2DM. Finally, we pointed out that other neuropsychiatric disorders, besides OCD, represent potential brain “insulinopathies”. Two distinct clusters of psychiatric disorders have emerged, in which the genetics of insulin-related traits/diseases may exert divergent pleiotropic effects: one consisting of AN, OCD, and SCZ, which showed negative genetic overlap with somatic insulin-related diseases and traits, and the other one comprising ADHD, and MDD, which showed positive genetic overlap with insulin-related diseases and traits. Our work might open up new directions for clinical and neuropsychopharmacological research by introducing insulin signalling as a possible mechanism underlying the multimorbidity of major mental and somatic illnesses. Further studies are warranted to investigate the biological meaning of the observed correlations and potential non-genetic effects contributing to insulin-related multimorbidity.

## Data Availability

The summary statistics of genome-wide association studies used as input for our analyses are available at the following links: for ADHD, AN, ASD, BIP, CROSS, OCD, MDD, TS - https://www.med.unc.edu/pgc/download-results/; AD - https://ctg.cncr.nl/software/summary_statistics; SCZ - http://walters.psycm.cf.ac.uk/; 2hGlu, FGlu, Fins, HbA1c, HOMA-IR - https://www.magicinvestigators.org/downloads/; BMI - https://portals.broadinstitute.org/collaboration/giant/index.php/GIANT_consortium_data_files.

## URLs

LDSC, https://github.com/bulik/ldsc; Pre-computed European LD scores, https://data.broadinstitute.org/alkesgroup/LDSCORE/; GNOVA, https://github.com/xtonyjiang/GNOVA; GWAS summary statistics -ADHD, AN, ASD, BIP, CROSS, OCD, MDD, TS: https://www.med.unc.edu/pgc/download-results/; AD: https://ctg.cncr.nl/software/summary_statistics; SCZ: http://walters.psycm.cf.ac.uk/; 2hGlu, FGlu, Fins, HbA1c, HOMA-IR: https://www.magicinvestigators.org/downloads/; BMI: https://portals.broadinstitute.org/collaboration/giant/index.php/GIANT_consortium_data_files; MSigDB: https://www.gsea-msigdb.org/gsea/msigdb/index.jsp.

## Acknowledgments

This project has received funding from the European Union’s Horizon 2020 research and innovation programme under grant agreement No 847879 (PRIME, Prevention and Remediation of Insulin Multimorbidity in Europe). NMR was supported by the European Union’s Horizon 2020 Programme (H2020/2014 – 2020) under grant agreement No 667302 (CoCA) and by funding for the Dutch National Science Agenda NeurolabNL project (grant 400-17-602). JB was supported by a personal grant from the Netherlands Organization for Scientific Research (NWO) Innovation Program (Veni grant No 09150161910091).

The analyses were carried out on the Dutch national e-infrastructure, part of the research programme Computing Time National Computing Facilities Processing Round pilots 2018 with project No 17666, which is (partly) financed by the NWO.

We also thank the researchers of the consortia that provided the GWAS summary statistics used in our analyses and the participants of the cohorts to which they refer.

