## Supplementary information for "Insulinopathies of the brain? Genetic overlap between somatic insulin-related and neuropsychiatric disorders"

1. **Genome-wide bivariate genetic correlation estimations**

First, the GWAS summary statistics were quality-controlled through the Linkage Disequilibrium Score Regression (LDSC) *munge_sumstats.py* function by removing single nucleotide polymorphisms (SNPs) that had a MAF≤0.01 or were poorly imputed (INFO score≤0.9 or not matching to HapMap3 SNPs, which tended to be well imputed in most studies). Strand-ambiguous variants, missing or out-of-bounds values (expected ranges for MAF: 0-1, INFO: 0-1.5, p-value: 0-1), and all indels were also removed. If the sample size varied from one SNP to the others, SNPs having an effective sample size [Neff = 4/(1/Cases + 1/Controls)] less than 0.67 times the 90^th^ percentile of sample size were removed. Pre-computed Linkage Disequilibrium (LD) scores, referring to the European population from the 1,000 Genomes Project (1kGP), were used as regression weights to estimate genetic correlations in LDSC. The major histocompatibility complex (MHC) locus (chr6:29-33 Mbp) was excluded from LD scores computation because of its high regional LD and the risk of producing bias in the regression. Standard errors and statistical significance were assessed via a block jack-knife procedure.

2. **Genetic covariance analyses stratified by functional annotations**

GeNetic cOVariance Analyzer (GNOVA) uses the method of moments as underlying framework to partition genetic covariances by functionally annotated subsets of SNPs. Insulin signalling-related gene-sets were identified and downloaded from Molecular Database v7.1 (MSigDB) (<https://www.gsea-msigdb.org/gsea/msigdb/index.jsp>) by using “insulin” as search keyword. Eight curated (by BioCarta, Broad Institute, Kyoto Encyclopedia of Genes and Genomes (KEGG), Massachusetts Institute of Technology, MSigDB Team, US National Institutes of Health, Nature publishing group, and Reactome) gene-sets were selected that relate directly to insulin signalling and have insulin in their name headers. We also considered an additional curated gene-set including 53 genes that are specifically relevant to central nervous system (CNS) insulin signalling, in addition to being listed among the top-ranked OCD GWAS findings or supported by animal/candidate gene/transcriptomic studies on OCD (1). Each gene-set was annotated to SNP positions on the 1kGP reference panel using the LDSC *make_annot.py* function. Then we performed stratified genetic covariance analyses of AD, ASD, obsessive-compulsive disorder (OCD) with metabolic syndrome (MetS), obesity and/or type 2 diabetes mellitus (T2DM) in GNOVA, using the LDSC-munged GWAS summary statistics of the considered phenotypes as input datasets, as well as the annotated SNP positions, as indicated in <https://github.com/xtonyjiang/GNOVA>.

3. **Samples characteristics**

Below, we report a summary of the characteristics of each genome-wide association study (GWAS), whose summary results were used in our analyses. For additional details about the GWAS methodology, please refer to the original publication, whose reference is also provided here. For each GWAS dataset, **Table 1** lists the number of cases and controls and the N_eff_ (only for binary phenotypes).

3.1. ***Samples included in the analyses***

**Obesity** (2)

A population-based case-control sample of European ethnicity from UK Biobank, release 2, was used for this GWAS. Three independent genome-wide loci associated with obesity were identified. Cases were identified by referring to UK Biobank field 41204 (“Diagnoses - secondary ICD-10 (International Classification of Diseases, 10th revision”) and phenotype code “E66 Overweight and obesity”.

**Metabolic syndrome (MetS)** (3)

A population-based case-control sample of European ethnicity from UK Biobank was used for this GWAS. Ninety-three independent genome-wide loci associated with MetS were identified. The National Cholesterol Education Program Expert Panel (NCEP) criteria for MetS were adopted to identify the five components of the syndrome. Three out of the following five criteria had to be met: high-density lipoprotein (HDL) cholesterol <1.0 mmol/L in men and <1.3 mmol/L in women, serum triglycerides ≥1.7 mmol/L, serum glucose ≥6.1 mmol/L or undergoing antidiabetic treatment, blood pressure ≥130/85 mmHg or undergoing antihypertensive treatment, waist circumference >102 cm in men and >88 cm in women.

**Type 2 diabetes mellitus (T2DM)** (4)

GWAS results for 32 studies, including individuals of European descendent, were aggregated through this case-control meta-analysis. Two hundred and forty-three risk loci associated with T2DM were identified. The T2DM diagnosis was obtained in accordance with the World Health Organisation (WHO) 1999 criteria or ICD-9 codes, based on hospital discharge diagnoses or electronic health records. The phenotype was defined based on a fasting glucose history ≥7.0 mmol/l or a 2-hour glucose ≥11.1 mmol/l during an oral glucose tolerance test (OGTT) or HbA1c ≥6.5% or use of antidiabetic drugs. Alternatively, in UK Biobank the phenotype was based on self-reported diabetes confirmed by an additional validated questionnaire. In order to prevent mixing with type 1 diabetes mellitus (T1DM), all patients were over 35 years of age at onset and had no detectable antibodies to glutamic acid decarboxylase (anti-GAD) or fasting C-peptide ≤0.30 nmol/L. Individuals were also excluded if they had known or probable first-degree familiarity for T1DM, or whether insulin treatment was started within ten years after the diagnosis.

**Alzheimer’s disease (AD)** (5)

The GWAS summary statistics used referred to the results of the Phase 3 analysis included in the Jansen and colleagues' work. In this phase, data from four cohorts, including individuals of European ancestry, were meta-analysed. The four cohorts were the case-control cohort of the Alzheimer's working group of the Psychiatric Genomic Consortium (PGC-ALZ), the International Genomics of Alzheimer's Project (IGAP) and the Alzheimer's Disease Sequencing Project (ADSP), and the population-based UK Biobank sample. Twentynine independent risk loci were identified. The diagnosis of AD followed the recommendations of the National Institute on Aging-Alzheimer's Association (NIA/AA), the NINCDS-ADRDA criteria, the ICD-10 research criteria or it was obtained from the National Patient Register, the Causes of Death Register, or the prescribed medication register. The raw data from the UK Biobank sample was used to create a proxy weighted AD phenotype. In detail, in UK Biobank the phenotype was constructed as a linear count of the number of affected biological parents (0, 1 or 2). The contribution of each non-affected parent to this count was weighted according to the age/age of the parent at the time of death to take into account the fact that they may not yet have passed through the risk period for this late-onset disease. Participants with a diagnosis of “Alzheimer's disease” (code G30) or “Dementia in Alzheimer's disease” (chapter on mental and behavioural disorders; code F00) as the cause of death or from previous hospitalisation records were treated as AD cases and obtained the highest possible risk score of 2, regardless of pathological family history. Due to the small number of the latter cases with a direct diagnosis in UK biobank (393 in all), this information was used to complement the proxy parent phenotype instead of as a primary outcome.

**Autism spectrum disorder (ASD)** (6)

Five family-based cohorts (i.e., the Geschwind Autism Center of Excellence (ACE), the Autism Genome Project (AGP), the Autism Genetic Resource Exchange (AGRE), the US National Institute of Mental Health (NIMH) Repository, the Montreal/Boston Collection (MONBOS), and the Simons Simplex Collection) of European ancestry as well as the Lundbeck Foundation Initiative for Integrative Psychiatric Research” - iPSYCH Danish population-based case-control sample were meta-analysed. Five loci were genome-wide associated with ASD after the meta-analysis. With regard to the included family-based studies, the diagnosis was formulated using usual research tools and consensus diagnoses by clinicians. In the population-based cohort, cases were identified through the Danish Central Psychiatric Research Registry and diagnosed by a psychiatrist according to ICD-10 criteria for atypical autism (F84.1), childhood autism (F84.0), Asperger's syndrome (F84.5), pervasive developmental disorder, unspecified (F84.9), and other pervasive developmental disorders (F84.8).

**Obsessive-compulsive disorder (OCD)** (7)

In this meta-analysis, two case-control and parent-offspring trios cohorts (International Obsessive Compulsive Disorder Foundation Genetics Collaborative (IOCDF-GC), and OCD Collaborative Genetics Association Studies (OCGAS)), including individuals of European ancestry, were combined. No locus was found to be genome-wide associated with OCD. The diagnosis was obtained in compliance with the DSM-IV (Diagnostic and Statistical Manual of Mental Disorders, 4th edition) criteria.

**Body mass index (BMI)** (8)

Data from the population-based UK Biobank -release 2 cohort, which includes individuals of European origin, where used for this GWAS. The phenotype considered was a continuous measure of BMI. Eight hundred and ninety-eight independent risk loci were identified.

**Glucose levels 2 hours after an oral glucose challenge (2hGlu)** (9)

Summary data for 2hGlu are derived from a meta-analysis of nine GWAS studies and a follow-up of 29 independent loci in 17 other studies, all including non-diabetic individuals of European ethnicity. The 2hGlu measurements were untransformed and adjusted for age, sex, BMI, and study-specific covariates. Individuals with diabetes (identified by previous diagnosis, ongoing antidiabetic treatment or fasting plasma glucose ≥7 mmol/L) were excluded from the study.

**Fasting glucose (FGlu) and insulin (FIns)** (10)

GWAS meta-analyses on fasting glucose levels (FGlu) measured in mmol/L and fasting insulin concentrations (FIns) measured in pmol/L were conducted in individuals of European ancestry. Data for FGlu and FIns were analysed without adjusting for BMI. Individuals with a medical diagnosis of diabetes, or treated with oral antidiabetic medications or insulin, or having a FGlu ≥7 mmol/L were excluded from the analyses. Some individual studies applied additional sample exclusions, such as pregnancy status, non-fasting individuals, and diagnosis of T1DM.

**Glycated haemoglobin (HbA1c)** (11)

Fifty-six population-based, prospective, case-control, or twin GWAS studies, including individuals of European descent, were meta-analysed. Forty-three independent loci were identified in European individuals. All participants were diabetes-free (as defined by physician diagnosis, medication use, or FGlu ≥7 mmol/L). When FGlu was not available, some single studies also eliminated subjects having 2hGlu ≥11.1 mmol/L, or HbA1c ≥6.5%.

**Homeostatic model assessment for insulin resistance (HOMA-IR)** (12)

Data from 20 GWAS, whose participants were all adults of European ancestry, were meta-analysed. Individuals were excluded from the analyses if they were diagnosed with T2DM, were being treated with oral anti-diabetic drugs or insulin or had FGlu ≥7 mmol/L. Individual studies applied additional sample exclusions, including pregnancy, non-fasting individuals, T1DM, or outliers ±3 standard deviations of the distribution for FGlu or FIns.

**Attention-deficit/hyperactivity disorder (ADHD)** (13)

The GWAS summary data were referred to a European subsample from the meta-analysis on ADHD, which was conducted on samples from the Psychiatric Genomics Consortium (PGC) and iPSYCH. The overall analysis, which considered 12 cohorts, revealed 12 loci with ADHD. For the iPSYCH cohort, cases were identified using the Danish Psychiatric Central Research Register and diagnoses were conﬁrmed by expert clinicians in accordance with ICD-10. The PGC cohorts included seven case-control and four family-based cohorts. Patients with ADHD were recruited from hospitals and clinics or by making use of medical registers. Diagnoses were made by trained personnel using research diagnostic tools.

**Anorexia nervosa (AN)** (14)

Thirty-three GWAS studies, including European individuals from the Anorexia Nervosa Genetics Initiative (ANGI), the Eating Disorders Working Group of the Psychiatric Genomics Consortium (PGC-ED) and UK Biobank cohorts as well as additional controls from Poland were combined in a meta-analysis on AN. Eight significant loci were identified. The AN phenotype was defined by referring to medical records from hospitals and other registries, structured clinical interviews or online questionnaires based on standardised diagnostic criteria (from the DSM-III-R, DSM-IV, ICD-8, ICD-9, ICD-10), or self-reported diagnoses for UK Biobank participants.

**Bipolar disorder (BIP)** (15)

Thirty-two case-control studies from Australia, North America, and Europe, including individuals of European descent, were meta-analysed. Thirty independent loci were genome-wide associated with BIP. The diagnosis of bipolar disorder was formulated according to the criteria indicated by the DSM-IV, ICD-9, or ICD-10, through the use of structured clinical interviews, medically administered checklists, or a review of medical records.

**Major depressive disorder (MDD)** (16, 17)

Summary data were derived from a GWAS meta-analysis of European individuals from the 33 cohorts of the Psychiatric Genomics Consortium (excluding UK Biobank and 23andMe data) as described in Wray et al., 2018 (16) and the broad depression phenotype in the full release of the UK Biobank as described in Howard et al., 2018 (17). The overall meta-analysis identified one-hundred and one independent loci. With regard to the PGC cohorts, cases were identified using structured diagnostic interviews (referring to standardised clinical criteria from DSM-V, ICD-9 and ICD-10), or review of electronic health records. In UK Biobank, the broad depression phenotype was obtained using self-reported help-seeking behaviour for mental health difﬁculties and affirmative response at least to one of the following two questions: “Have you ever seen a general practitioner (GP) for nerves, anxiety, tension or depression?” (ﬁeld 2090) or “Have you ever seen a psychiatrist for nerves, anxiety, tension or depression?” (ﬁeld 2010); alternatively, primary or secondary diagnoses of a depressive mood disorder from hospital medical records were used (ﬁelds 41202 and 41204; ICD codes: F32—Single Episode Depression, F33—Recurrent Depression, F34—Persistent mood disorders, F38—Other mood disorders and F39—Unspeciﬁed mood disorders).

**Schizophrenia (SCZ)** (18)

The summary data were referred to a meta-analysis of SCZ GWAS data from 46 case-control cohorts of European descendent. In this GWAS meta-analysis, PGC data were reanalysed by including a larger CLOZUK subsample. The meta-analysis identified one-hundred and forty-five loci associated with SCZ. Cases included patients either with SCZ or schizoaffective disorder, and they were identified by clinical diagnosis or using research-based assessment tools, depending on the cohort.

**Tourette’s syndrome (TS)** (19)

The GWAS meta-analysis on TS included three case-control cohorts and one family-based cohort from Europe and North America, including individuals of European. One genome-wide significant locus was associated with TS. Most of the cases were diagnosed in accordance with DSM-IV-TR or DSM-V criteria for TS. Twelve cases met DSM-V criteria for chronic vocal or motor tic disorder. All cases were recruited in specialised clinics or by online recruitment combined with web-based, validated phenotypic assessments.

9. Saxena R, Hivert MF, Langenberg C, Tanaka T, Pankow JS, Vollenweider P, Lyssenko V, Bouatia-Naji N, Dupuis J, Jackson AU, Kao WH, Li M, Glazer NL, Manning AK, Luan J, Stringham HM, Prokopenko I, Johnson T, Grarup N, Boesgaard TW, Lecoeur C, Shrader P, O'Connell J, Ingelsson E, Couper DJ, Rice K, Song K, Andreasen CH, Dina C, Kottgen A, Le Bacquer O, Pattou F, Taneera J, Steinthorsdottir V, Rybin D, Ardlie K, Sampson M, Qi L, van Hoek M, Weedon MN, Aulchenko YS, Voight BF, Grallert H, Balkau B, Bergman RN, Bielinski SJ, Bonnefond A, Bonnycastle LL, Borch-Johnsen K, Bottcher Y, Brunner E, Buchanan TA, Bumpstead SJ, Cavalcanti-Proenca C, Charpentier G, Chen YD, Chines PS, Collins FS, Cornelis M, G JC, Delplanque J, Doney A, Egan JM, Erdos MR, Firmann M, Forouhi NG, Fox CS, Goodarzi MO, Graessler J, Hingorani A, Isomaa B, Jorgensen T, Kivimaki M, Kovacs P, Krohn K, Kumari M, Lauritzen T, Levy-Marchal C, Mayor V, McAteer JB, Meyre D, Mitchell BD, Mohlke KL, Morken MA, Narisu N, Palmer CN, Pakyz R, Pascoe L, Payne F, Pearson D, Rathmann W, Sandbaek A, Sayer AA, Scott LJ, Sharp SJ, Sijbrands E, Singleton A, Siscovick DS, Smith NL, Sparso T, Swift AJ, Syddall H, Thorleifsson G, Tonjes A, Tuomi T, Tuomilehto J, Valle TT, Waeber G, Walley A, Waterworth DM, Zeggini E, Zhao JH, consortium G, investigators M, Illig T, Wichmann HE, Wilson JF, van Duijn C, Hu FB, Morris AD, Frayling TM, Hattersley AT, Thorsteinsdottir U, Stefansson K, Nilsson P, Syvanen AC, Shuldiner AR, Walker M, Bornstein SR, Schwarz P, Williams GH, Nathan DM, Kuusisto J, Laakso M, Cooper C, Marmot M, Ferrucci L, Mooser V, Stumvoll M, Loos RJ, Altshuler D, Psaty BM, Rotter JI, Boerwinkle E, Hansen T, Pedersen O, Florez JC, McCarthy MI, Boehnke M, Barroso I, Sladek R, Froguel P, Meigs JB, Groop L, Wareham NJ, Watanabe RM. Genetic variation in GIPR influences the glucose and insulin responses to an oral glucose challenge. Nat Genet. 2010;42:142-148.

10. Lagou V, Magi R, Hottenga JJ, Grallert H, Perry JRB, Bouatia-Naji N, Marullo L, Rybin D, Jansen R, Min JL, Dimas AS, Ulrich A, Zudina L, Gadin JR, Jiang L, Faggian A, Bonnefond A, Fadista J, Stathopoulou MG, Isaacs A, Willems SM, Navarro P, Tanaka T, Jackson AU, Montasser ME, O'Connell JR, Bielak LF, Webster RJ, Saxena R, Stafford JM, Pourcain BS, Timpson NJ, Salo P, Shin SY, Amin N, Smith AV, Li G, Verweij N, Goel A, Ford I, Johnson PCD, Johnson T, Kapur K, Thorleifsson G, Strawbridge RJ, Rasmussen-Torvik LJ, Esko T, Mihailov E, Fall T, Fraser RM, Mahajan A, Kanoni S, Giedraitis V, Kleber ME, Silbernagel G, Meyer J, Muller-Nurasyid M, Ganna A, Sarin AP, Yengo L, Shungin D, Luan J, Horikoshi M, An P, Sanna S, Boettcher Y, Rayner NW, Nolte IM, Zemunik T, Iperen EV, Kovacs P, Hastie ND, Wild SH, McLachlan S, Campbell S, Polasek O, Carlson O, Egan J, Kiess W, Willemsen G, Kuusisto J, Laakso M, Dimitriou M, Hicks AA, Rauramaa R, Bandinelli S, Thorand B, Liu Y, Miljkovic I, Lind L, Doney A, Perola M, Hingorani A, Kivimaki M, Kumari M, Bennett AJ, Groves CJ, Herder C, Koistinen HA, Kinnunen L, Faire U, Bakker SJL, Uusitupa M, Palmer CNA, Jukema JW, Sattar N, Pouta A, Snieder H, Boerwinkle E, Pankow JS, Magnusson PK, Krus U, Scapoli C, de Geus E, Bluher M, Wolffenbuttel BHR, Province MA, Abecasis GR, Meigs JB, Hovingh GK, Lindstrom J, Wilson JF, Wright AF, Dedoussis GV, Bornstein SR, Schwarz PEH, Tonjes A, Winkelmann BR, Boehm BO, Marz W, Metspalu A, Price JF, Deloukas P, Korner A, Lakka TA, Keinanen-Kiukaanniemi SM, Saaristo TE, Bergman RN, Tuomilehto J, Wareham NJ, Langenberg C, Mannisto S, Franks PW, Hayward C, Vitart V, Kaprio J, Visvikis-Siest S, Balkau B, Altshuler D, Rudan I, Stumvoll M, Campbell H, van Duijn CM, Gieger C, Illig T, Ferrucci L, Pedersen NL, Pramstaller PP, Boehnke M, Frayling TM, Shuldiner AR, Peyser PA, Kardia SLR, Palmer LJ, Penninx BW, Meneton P, Harris TB, Navis G, Harst PV, Smith GD, Forouhi NG, Loos RJF, Salomaa V, Soranzo N, Boomsma DI, Groop L, Tuomi T, Hofman A, Munroe PB, Gudnason V, Siscovick DS, Watkins H, Lecoeur C, Vollenweider P, Franco-Cereceda A, Eriksson P, Jarvelin MR, Stefansson K, Hamsten A, Nicholson G, Karpe F, Dermitzakis ET, Lindgren CM, McCarthy MI, Froguel P, Kaakinen MA, Lyssenko V, Watanabe RM, Ingelsson E, Florez JC, Dupuis J, Barroso I, Morris AP, Prokopenko I, Meta-Analyses of G, Insulin-related traits C. Sex-dimorphic genetic effects and novel loci for fasting glucose and insulin variability. Nat Commun. 2021;12:24.

11. Wheeler E, Leong A, Liu CT, Hivert MF, Strawbridge RJ, Podmore C, Li M, Yao J, Sim X, Hong J, Chu AY, Zhang W, Wang X, Chen P, Maruthur NM, Porneala BC, Sharp SJ, Jia Y, Kabagambe EK, Chang LC, Chen WM, Elks CE, Evans DS, Fan Q, Giulianini F, Go MJ, Hottenga JJ, Hu Y, Jackson AU, Kanoni S, Kim YJ, Kleber ME, Ladenvall C, Lecoeur C, Lim SH, Lu Y, Mahajan A, Marzi C, Nalls MA, Navarro P, Nolte IM, Rose LM, Rybin DV, Sanna S, Shi Y, Stram DO, Takeuchi F, Tan SP, van der Most PJ, Van Vliet-Ostaptchouk JV, Wong A, Yengo L, Zhao W, Goel A, Martinez Larrad MT, Radke D, Salo P, Tanaka T, van Iperen EPA, Abecasis G, Afaq S, Alizadeh BZ, Bertoni AG, Bonnefond A, Bottcher Y, Bottinger EP, Campbell H, Carlson OD, Chen CH, Cho YS, Garvey WT, Gieger C, Goodarzi MO, Grallert H, Hamsten A, Hartman CA, Herder C, Hsiung CA, Huang J, Igase M, Isono M, Katsuya T, Khor CC, Kiess W, Kohara K, Kovacs P, Lee J, Lee WJ, Lehne B, Li H, Liu J, Lobbens S, Luan J, Lyssenko V, Meitinger T, Miki T, Miljkovic I, Moon S, Mulas A, Muller G, Muller-Nurasyid M, Nagaraja R, Nauck M, Pankow JS, Polasek O, Prokopenko I, Ramos PS, Rasmussen-Torvik L, Rathmann W, Rich SS, Robertson NR, Roden M, Roussel R, Rudan I, Scott RA, Scott WR, Sennblad B, Siscovick DS, Strauch K, Sun L, Swertz M, Tajuddin SM, Taylor KD, Teo YY, Tham YC, Tonjes A, Wareham NJ, Willemsen G, Wilsgaard T, Hingorani AD, Consortium E-C, Consortium EP-I, Lifelines Cohort S, Egan J, Ferrucci L, Hovingh GK, Jula A, Kivimaki M, Kumari M, Njolstad I, Palmer CNA, Serrano Rios M, Stumvoll M, Watkins H, Aung T, Bluher M, Boehnke M, Boomsma DI, Bornstein SR, Chambers JC, Chasman DI, Chen YI, Chen YT, Cheng CY, Cucca F, de Geus EJC, Deloukas P, Evans MK, Fornage M, Friedlander Y, Froguel P, Groop L, Gross MD, Harris TB, Hayward C, Heng CK, Ingelsson E, Kato N, Kim BJ, Koh WP, Kooner JS, Korner A, Kuh D, Kuusisto J, Laakso M, Lin X, Liu Y, Loos RJF, Magnusson PKE, Marz W, McCarthy MI, Oldehinkel AJ, Ong KK, Pedersen NL, Pereira MA, Peters A, Ridker PM, Sabanayagam C, Sale M, Saleheen D, Saltevo J, Schwarz PE, Sheu WHH, Snieder H, Spector TD, Tabara Y, Tuomilehto J, van Dam RM, Wilson JG, Wilson JF, Wolffenbuttel BHR, Wong TY, Wu JY, Yuan JM, Zonderman AB, Soranzo N, Guo X, Roberts DJ, Florez JC, Sladek R, Dupuis J, Morris AP, Tai ES, Selvin E, Rotter JI, Langenberg C, Barroso I, Meigs JB. Impact of common genetic determinants of Hemoglobin A1c on type 2 diabetes risk and diagnosis in ancestrally diverse populations: A transethnic genome-wide meta-analysis. PLoS Med. 2017;14:e1002383.

12. Dupuis J, Langenberg C, Prokopenko I, Saxena R, Soranzo N, Jackson AU, Wheeler E, Glazer NL, Bouatia-Naji N, Gloyn AL, Lindgren CM, Magi R, Morris AP, Randall J, Johnson T, Elliott P, Rybin D, Thorleifsson G, Steinthorsdottir V, Henneman P, Grallert H, Dehghan A, Hottenga JJ, Franklin CS, Navarro P, Song K, Goel A, Perry JR, Egan JM, Lajunen T, Grarup N, Sparso T, Doney A, Voight BF, Stringham HM, Li M, Kanoni S, Shrader P, Cavalcanti-Proenca C, Kumari M, Qi L, Timpson NJ, Gieger C, Zabena C, Rocheleau G, Ingelsson E, An P, O'Connell J, Luan J, Elliott A, McCarroll SA, Payne F, Roccasecca RM, Pattou F, Sethupathy P, Ardlie K, Ariyurek Y, Balkau B, Barter P, Beilby JP, Ben-Shlomo Y, Benediktsson R, Bennett AJ, Bergmann S, Bochud M, Boerwinkle E, Bonnefond A, Bonnycastle LL, Borch-Johnsen K, Bottcher Y, Brunner E, Bumpstead SJ, Charpentier G, Chen YD, Chines P, Clarke R, Coin LJ, Cooper MN, Cornelis M, Crawford G, Crisponi L, Day IN, de Geus EJ, Delplanque J, Dina C, Erdos MR, Fedson AC, Fischer-Rosinsky A, Forouhi NG, Fox CS, Frants R, Franzosi MG, Galan P, Goodarzi MO, Graessler J, Groves CJ, Grundy S, Gwilliam R, Gyllensten U, Hadjadj S, Hallmans G, Hammond N, Han X, Hartikainen AL, Hassanali N, Hayward C, Heath SC, Hercberg S, Herder C, Hicks AA, Hillman DR, Hingorani AD, Hofman A, Hui J, Hung J, Isomaa B, Johnson PR, Jorgensen T, Jula A, Kaakinen M, Kaprio J, Kesaniemi YA, Kivimaki M, Knight B, Koskinen S, Kovacs P, Kyvik KO, Lathrop GM, Lawlor DA, Le Bacquer O, Lecoeur C, Li Y, Lyssenko V, Mahley R, Mangino M, Manning AK, Martinez-Larrad MT, McAteer JB, McCulloch LJ, McPherson R, Meisinger C, Melzer D, Meyre D, Mitchell BD, Morken MA, Mukherjee S, Naitza S, Narisu N, Neville MJ, Oostra BA, Orru M, Pakyz R, Palmer CN, Paolisso G, Pattaro C, Pearson D, Peden JF, Pedersen NL, Perola M, Pfeiffer AF, Pichler I, Polasek O, Posthuma D, Potter SC, Pouta A, Province MA, Psaty BM, Rathmann W, Rayner NW, Rice K, Ripatti S, Rivadeneira F, Roden M, Rolandsson O, Sandbaek A, Sandhu M, Sanna S, Sayer AA, Scheet P, Scott LJ, Seedorf U, Sharp SJ, Shields B, Sigurethsson G, Sijbrands EJ, Silveira A, Simpson L, Singleton A, Smith NL, Sovio U, Swift A, Syddall H, Syvanen AC, Tanaka T, Thorand B, Tichet J, Tonjes A, Tuomi T, Uitterlinden AG, van Dijk KW, van Hoek M, Varma D, Visvikis-Siest S, Vitart V, Vogelzangs N, Waeber G, Wagner PJ, Walley A, Walters GB, Ward KL, Watkins H, Weedon MN, Wild SH, Willemsen G, Witteman JC, Yarnell JW, Zeggini E, Zelenika D, Zethelius B, Zhai G, Zhao JH, Zillikens MC, Consortium D, Consortium G, Global BC, Borecki IB, Loos RJ, Meneton P, Magnusson PK, Nathan DM, Williams GH, Hattersley AT, Silander K, Salomaa V, Smith GD, Bornstein SR, Schwarz P, Spranger J, Karpe F, Shuldiner AR, Cooper C, Dedoussis GV, Serrano-Rios M, Morris AD, Lind L, Palmer LJ, Hu FB, Franks PW, Ebrahim S, Marmot M, Kao WH, Pankow JS, Sampson MJ, Kuusisto J, Laakso M, Hansen T, Pedersen O, Pramstaller PP, Wichmann HE, Illig T, Rudan I, Wright AF, Stumvoll M, Campbell H, Wilson JF, Anders Hamsten on behalf of Procardis C, investigators M, Bergman RN, Buchanan TA, Collins FS, Mohlke KL, Tuomilehto J, Valle TT, Altshuler D, Rotter JI, Siscovick DS, Penninx BW, Boomsma DI, Deloukas P, Spector TD, Frayling TM, Ferrucci L, Kong A, Thorsteinsdottir U, Stefansson K, van Duijn CM, Aulchenko YS, Cao A, Scuteri A, Schlessinger D, Uda M, Ruokonen A, Jarvelin MR, Waterworth DM, Vollenweider P, Peltonen L, Mooser V, Abecasis GR, Wareham NJ, Sladek R, Froguel P, Watanabe RM, Meigs JB, Groop L, Boehnke M, McCarthy MI, Florez JC, Barroso I. New genetic loci implicated in fasting glucose homeostasis and their impact on type 2 diabetes risk. Nat Genet. 2010;42:105-116.

13. Demontis D, Walters RK, Martin J, Mattheisen M, Als TD, Agerbo E, Baldursson G, Belliveau R, Bybjerg-Grauholm J, Baekvad-Hansen M, Cerrato F, Chambert K, Churchhouse C, Dumont A, Eriksson N, Gandal M, Goldstein JI, Grasby KL, Grove J, Gudmundsson OO, Hansen CS, Hauberg ME, Hollegaard MV, Howrigan DP, Huang H, Maller JB, Martin AR, Martin NG, Moran J, Pallesen J, Palmer DS, Pedersen CB, Pedersen MG, Poterba T, Poulsen JB, Ripke S, Robinson EB, Satterstrom FK, Stefansson H, Stevens C, Turley P, Walters GB, Won H, Wright MJ, Consortium AWGotPG, Early L, Genetic Epidemiology C, andMe Research T, Andreassen OA, Asherson P, Burton CL, Boomsma DI, Cormand B, Dalsgaard S, Franke B, Gelernter J, Geschwind D, Hakonarson H, Haavik J, Kranzler HR, Kuntsi J, Langley K, Lesch KP, Middeldorp C, Reif A, Rohde LA, Roussos P, Schachar R, Sklar P, Sonuga-Barke EJS, Sullivan PF, Thapar A, Tung JY, Waldman ID, Medland SE, Stefansson K, Nordentoft M, Hougaard DM, Werge T, Mors O, Mortensen PB, Daly MJ, Faraone SV, Borglum AD, Neale BM. Discovery of the first genome-wide significant risk loci for attention deficit/hyperactivity disorder. Nat Genet. 2019;51:63-75.

14. Watson HJ, Yilmaz Z, Thornton LM, Hubel C, Coleman JRI, Gaspar HA, Bryois J, Hinney A, Leppa VM, Mattheisen M, Medland SE, Ripke S, Yao S, Giusti-Rodriguez P, Anorexia Nervosa Genetics I, Hanscombe KB, Purves KL, Eating Disorders Working Group of the Psychiatric Genomics C, Adan RAH, Alfredsson L, Ando T, Andreassen OA, Baker JH, Berrettini WH, Boehm I, Boni C, Perica VB, Buehren K, Burghardt R, Cassina M, Cichon S, Clementi M, Cone RD, Courtet P, Crow S, Crowley JJ, Danner UN, Davis OSP, de Zwaan M, Dedoussis G, Degortes D, DeSocio JE, Dick DM, Dikeos D, Dina C, Dmitrzak-Weglarz M, Docampo E, Duncan LE, Egberts K, Ehrlich S, Escaramis G, Esko T, Estivill X, Farmer A, Favaro A, Fernandez-Aranda F, Fichter MM, Fischer K, Focker M, Foretova L, Forstner AJ, Forzan M, Franklin CS, Gallinger S, Giegling I, Giuranna J, Gonidakis F, Gorwood P, Mayora MG, Guillaume S, Guo Y, Hakonarson H, Hatzikotoulas K, Hauser J, Hebebrand J, Helder SG, Herms S, Herpertz-Dahlmann B, Herzog W, Huckins LM, Hudson JI, Imgart H, Inoko H, Janout V, Jimenez-Murcia S, Julia A, Kalsi G, Kaminska D, Kaprio J, Karhunen L, Karwautz A, Kas MJH, Kennedy JL, Keski-Rahkonen A, Kiezebrink K, Kim YR, Klareskog L, Klump KL, Knudsen GPS, La Via MC, Le Hellard S, Levitan RD, Li D, Lilenfeld L, Lin BD, Lissowska J, Luykx J, Magistretti PJ, Maj M, Mannik K, Marsal S, Marshall CR, Mattingsdal M, McDevitt S, McGuffin P, Metspalu A, Meulenbelt I, Micali N, Mitchell K, Monteleone AM, Monteleone P, Munn-Chernoff MA, Nacmias B, Navratilova M, Ntalla I, O'Toole JK, Ophoff RA, Padyukov L, Palotie A, Pantel J, Papezova H, Pinto D, Rabionet R, Raevuori A, Ramoz N, Reichborn-Kjennerud T, Ricca V, Ripatti S, Ritschel F, Roberts M, Rotondo A, Rujescu D, Rybakowski F, Santonastaso P, Scherag A, Scherer SW, Schmidt U, Schork NJ, Schosser A, Seitz J, Slachtova L, Slagboom PE, Slof-Op 't Landt MCT, Slopien A, Sorbi S, Swiatkowska B, Szatkiewicz JP, Tachmazidou I, Tenconi E, Tortorella A, Tozzi F, Treasure J, Tsitsika A, Tyszkiewicz-Nwafor M, Tziouvas K, van Elburg AA, van Furth EF, Wagner G, Walton E, Widen E, Zeggini E, Zerwas S, Zipfel S, Bergen AW, Boden JM, Brandt H, Crawford S, Halmi KA, Horwood LJ, Johnson C, Kaplan AS, Kaye WH, Mitchell JE, Olsen CM, Pearson JF, Pedersen NL, Strober M, Werge T, Whiteman DC, Woodside DB, Stuber GD, Gordon S, Grove J, Henders AK, Jureus A, Kirk KM, Larsen JT, Parker R, Petersen L, Jordan J, Kennedy M, Montgomery GW, Wade TD, Birgegard A, Lichtenstein P, Norring C, Landen M, Martin NG, Mortensen PB, Sullivan PF, Breen G, Bulik CM. Genome-wide association study identifies eight risk loci and implicates metabo-psychiatric origins for anorexia nervosa. Nat Genet. 2019;51:1207-1214.

15. Stahl EA, Breen G, Forstner AJ, McQuillin A, Ripke S, Trubetskoy V, Mattheisen M, Wang Y, Coleman JRI, Gaspar HA, de Leeuw CA, Steinberg S, Pavlides JMW, Trzaskowski M, Byrne EM, Pers TH, Holmans PA, Richards AL, Abbott L, Agerbo E, Akil H, Albani D, Alliey-Rodriguez N, Als TD, Anjorin A, Antilla V, Awasthi S, Badner JA, Baekvad-Hansen M, Barchas JD, Bass N, Bauer M, Belliveau R, Bergen SE, Pedersen CB, Boen E, Boks MP, Boocock J, Budde M, Bunney W, Burmeister M, Bybjerg-Grauholm J, Byerley W, Casas M, Cerrato F, Cervantes P, Chambert K, Charney AW, Chen D, Churchhouse C, Clarke TK, Coryell W, Craig DW, Cruceanu C, Curtis D, Czerski PM, Dale AM, de Jong S, Degenhardt F, Del-Favero J, DePaulo JR, Djurovic S, Dobbyn AL, Dumont A, Elvsashagen T, Escott-Price V, Fan CC, Fischer SB, Flickinger M, Foroud TM, Forty L, Frank J, Fraser C, Freimer NB, Frisen L, Gade K, Gage D, Garnham J, Giambartolomei C, Pedersen MG, Goldstein J, Gordon SD, Gordon-Smith K, Green EK, Green MJ, Greenwood TA, Grove J, Guan W, Guzman-Parra J, Hamshere ML, Hautzinger M, Heilbronner U, Herms S, Hipolito M, Hoffmann P, Holland D, Huckins L, Jamain S, Johnson JS, Jureus A, Kandaswamy R, Karlsson R, Kennedy JL, Kittel-Schneider S, Knowles JA, Kogevinas M, Koller AC, Kupka R, Lavebratt C, Lawrence J, Lawson WB, Leber M, Lee PH, Levy SE, Li JZ, Liu C, Lucae S, Maaser A, MacIntyre DJ, Mahon PB, Maier W, Martinsson L, McCarroll S, McGuffin P, McInnis MG, McKay JD, Medeiros H, Medland SE, Meng F, Milani L, Montgomery GW, Morris DW, Muhleisen TW, Mullins N, Nguyen H, Nievergelt CM, Adolfsson AN, Nwulia EA, O'Donovan C, Loohuis LMO, Ori APS, Oruc L, Osby U, Perlis RH, Perry A, Pfennig A, Potash JB, Purcell SM, Regeer EJ, Reif A, Reinbold CS, Rice JP, Rivas F, Rivera M, Roussos P, Ruderfer DM, Ryu E, Sanchez-Mora C, Schatzberg AF, Scheftner WA, Schork NJ, Shannon Weickert C, Shehktman T, Shilling PD, Sigurdsson E, Slaney C, Smeland OB, Sobell JL, Soholm Hansen C, Spijker AT, St Clair D, Steffens M, Strauss JS, Streit F, Strohmaier J, Szelinger S, Thompson RC, Thorgeirsson TE, Treutlein J, Vedder H, Wang W, Watson SJ, Weickert TW, Witt SH, Xi S, Xu W, Young AH, Zandi P, Zhang P, Zollner S, e QC, Consortium B, Adolfsson R, Agartz I, Alda M, Backlund L, Baune BT, Bellivier F, Berrettini WH, Biernacka JM, Blackwood DHR, Boehnke M, Borglum AD, Corvin A, Craddock N, Daly MJ, Dannlowski U, Esko T, Etain B, Frye M, Fullerton JM, Gershon ES, Gill M, Goes F, Grigoroiu-Serbanescu M, Hauser J, Hougaard DM, Hultman CM, Jones I, Jones LA, Kahn RS, Kirov G, Landen M, Leboyer M, Lewis CM, Li QS, Lissowska J, Martin NG, Mayoral F, McElroy SL, McIntosh AM, McMahon FJ, Melle I, Metspalu A, Mitchell PB, Morken G, Mors O, Mortensen PB, Muller-Myhsok B, Myers RM, Neale BM, Nimgaonkar V, Nordentoft M, Nothen MM, O'Donovan MC, Oedegaard KJ, Owen MJ, Paciga SA, Pato C, Pato MT, Posthuma D, Ramos-Quiroga JA, Ribases M, Rietschel M, Rouleau GA, Schalling M, Schofield PR, Schulze TG, Serretti A, Smoller JW, Stefansson H, Stefansson K, Stordal E, Sullivan PF, Turecki G, Vaaler AE, Vieta E, Vincent JB, Werge T, Nurnberger JI, Wray NR, Di Florio A, Edenberg HJ, Cichon S, Ophoff RA, Scott LJ, Andreassen OA, Kelsoe J, Sklar P, Bipolar Disorder Working Group of the Psychiatric Genomics C. Genome-wide association study identifies 30 loci associated with bipolar disorder. Nat Genet. 2019;51:793-803.

16. Wray NR, Ripke S, Mattheisen M, Trzaskowski M, Byrne EM, Abdellaoui A, Adams MJ, Agerbo E, Air TM, Andlauer TMF, Bacanu SA, Baekvad-Hansen M, Beekman AFT, Bigdeli TB, Binder EB, Blackwood DRH, Bryois J, Buttenschon HN, Bybjerg-Grauholm J, Cai N, Castelao E, Christensen JH, Clarke TK, Coleman JIR, Colodro-Conde L, Couvy-Duchesne B, Craddock N, Crawford GE, Crowley CA, Dashti HS, Davies G, Deary IJ, Degenhardt F, Derks EM, Direk N, Dolan CV, Dunn EC, Eley TC, Eriksson N, Escott-Price V, Kiadeh FHF, Finucane HK, Forstner AJ, Frank J, Gaspar HA, Gill M, Giusti-Rodriguez P, Goes FS, Gordon SD, Grove J, Hall LS, Hannon E, Hansen CS, Hansen TF, Herms S, Hickie IB, Hoffmann P, Homuth G, Horn C, Hottenga JJ, Hougaard DM, Hu M, Hyde CL, Ising M, Jansen R, Jin F, Jorgenson E, Knowles JA, Kohane IS, Kraft J, Kretzschmar WW, Krogh J, Kutalik Z, Lane JM, Li Y, Li Y, Lind PA, Liu X, Lu L, MacIntyre DJ, MacKinnon DF, Maier RM, Maier W, Marchini J, Mbarek H, McGrath P, McGuffin P, Medland SE, Mehta D, Middeldorp CM, Mihailov E, Milaneschi Y, Milani L, Mill J, Mondimore FM, Montgomery GW, Mostafavi S, Mullins N, Nauck M, Ng B, Nivard MG, Nyholt DR, O'Reilly PF, Oskarsson H, Owen MJ, Painter JN, Pedersen CB, Pedersen MG, Peterson RE, Pettersson E, Peyrot WJ, Pistis G, Posthuma D, Purcell SM, Quiroz JA, Qvist P, Rice JP, Riley BP, Rivera M, Saeed Mirza S, Saxena R, Schoevers R, Schulte EC, Shen L, Shi J, Shyn SI, Sigurdsson E, Sinnamon GBC, Smit JH, Smith DJ, Stefansson H, Steinberg S, Stockmeier CA, Streit F, Strohmaier J, Tansey KE, Teismann H, Teumer A, Thompson W, Thomson PA, Thorgeirsson TE, Tian C, Traylor M, Treutlein J, Trubetskoy V, Uitterlinden AG, Umbricht D, Van der Auwera S, van Hemert AM, Viktorin A, Visscher PM, Wang Y, Webb BT, Weinsheimer SM, Wellmann J, Willemsen G, Witt SH, Wu Y, Xi HS, Yang J, Zhang F, eQtlgen, andMe, Arolt V, Baune BT, Berger K, Boomsma DI, Cichon S, Dannlowski U, de Geus ECJ, DePaulo JR, Domenici E, Domschke K, Esko T, Grabe HJ, Hamilton SP, Hayward C, Heath AC, Hinds DA, Kendler KS, Kloiber S, Lewis G, Li QS, Lucae S, Madden PFA, Magnusson PK, Martin NG, McIntosh AM, Metspalu A, Mors O, Mortensen PB, Muller-Myhsok B, Nordentoft M, Nothen MM, O'Donovan MC, Paciga SA, Pedersen NL, Penninx B, Perlis RH, Porteous DJ, Potash JB, Preisig M, Rietschel M, Schaefer C, Schulze TG, Smoller JW, Stefansson K, Tiemeier H, Uher R, Volzke H, Weissman MM, Werge T, Winslow AR, Lewis CM, Levinson DF, Breen G, Borglum AD, Sullivan PF, Major Depressive Disorder Working Group of the Psychiatric Genomics C. Genome-wide association analyses identify 44 risk variants and refine the genetic architecture of major depression. Nat Genet. 2018;50:668-681.

17. Howard DM, Adams MJ, Shirali M, Clarke TK, Marioni RE, Davies G, Coleman JRI, Alloza C, Shen X, Barbu MC, Wigmore EM, Gibson J, andMe Research T, Hagenaars SP, Lewis CM, Ward J, Smith DJ, Sullivan PF, Haley CS, Breen G, Deary IJ, McIntosh AM. Genome-wide association study of depression phenotypes in UK Biobank identifies variants in excitatory synaptic pathways. Nat Commun. 2018;9:1470.

18. Pardinas AF, Holmans P, Pocklington AJ, Escott-Price V, Ripke S, Carrera N, Legge SE, Bishop S, Cameron D, Hamshere ML, Han J, Hubbard L, Lynham A, Mantripragada K, Rees E, MacCabe JH, McCarroll SA, Baune BT, Breen G, Byrne EM, Dannlowski U, Eley TC, Hayward C, Martin NG, McIntosh AM, Plomin R, Porteous DJ, Wray NR, Caballero A, Geschwind DH, Huckins LM, Ruderfer DM, Santiago E, Sklar P, Stahl EA, Won H, Agerbo E, Als TD, Andreassen OA, Baekvad-Hansen M, Mortensen PB, Pedersen CB, Borglum AD, Bybjerg-Grauholm J, Djurovic S, Durmishi N, Pedersen MG, Golimbet V, Grove J, Hougaard DM, Mattheisen M, Molden E, Mors O, Nordentoft M, Pejovic-Milovancevic M, Sigurdsson E, Silagadze T, Hansen CS, Stefansson K, Stefansson H, Steinberg S, Tosato S, Werge T, Consortium G, Consortium C, Collier DA, Rujescu D, Kirov G, Owen MJ, O'Donovan MC, Walters JTR. Common schizophrenia alleles are enriched in mutation-intolerant genes and in regions under strong background selection. Nat Genet. 2018;50:381-389.

19. Yu D, Sul JH, Tsetsos F, Nawaz MS, Huang AY, Zelaya I, Illmann C, Osiecki L, Darrow SM, Hirschtritt ME, Greenberg E, Muller-Vahl KR, Stuhrmann M, Dion Y, Rouleau G, Aschauer H, Stamenkovic M, Schlogelhofer M, Sandor P, Barr CL, Grados M, Singer HS, Nothen MM, Hebebrand J, Hinney A, King RA, Fernandez TV, Barta C, Tarnok Z, Nagy P, Depienne C, Worbe Y, Hartmann A, Budman CL, Rizzo R, Lyon GJ, McMahon WM, Batterson JR, Cath DC, Malaty IA, Okun MS, Berlin C, Woods DW, Lee PC, Jankovic J, Robertson MM, Gilbert DL, Brown LW, Coffey BJ, Dietrich A, Hoekstra PJ, Kuperman S, Zinner SH, Luethvigsson P, Saemundsen E, Thorarensen O, Atzmon G, Barzilai N, Wagner M, Moessner R, Ophoff R, Pato CN, Pato MT, Knowles JA, Roffman JL, Smoller JW, Buckner RL, Willsey AJ, Tischfield JA, Heiman GA, Stefansson H, Stefansson K, Posthuma D, Cox NJ, Pauls DL, Freimer NB, Neale BM, Davis LK, Paschou P, Coppola G, Mathews CA, Scharf JM, Tourette Association of America International Consortium for Genetics tGdlTGRItTICGS, the Psychiatric Genomics Consortium Tourette Syndrome Working G. Interrogating the Genetic Determinants of Tourette's Syndrome and Other Tic Disorders Through Genome-Wide Association Studies. Am J Psychiatry. 2019;176:217-227.
